# Effect of a male-targeted digital decision support application aimed at increasing linkage to HIV care among men: Findings from the HITS cluster randomized clinical trial in rural South Africa

**DOI:** 10.1101/2024.03.15.24304373

**Authors:** Hae-Young Kim, Maxime Inghels, Thulile Mathenjwa, Maryam Shahmanesh, Janet Seeley, Phillippa Matthews, Sally Wyke, Nuala McGrath, Oluwafemi Adeagbo, Dickman Gareta, H. Manisha Yapa, Thembelihle Zuma, Adrian Dobra, Ann Blandford, Till Bärnighausen, Frank Tanser

**Author notes:** <u>Corresponding Author:</u> Hae-Young Kim, PhD, 227 East 30^th^ Street, New York, NY, USA. **Competing interests** The authors declare no conflicts of interest. **Authors’ contributions** **Oluwafemi Adeagbo:** Conceptualization, Methodology, Software, Writing – Review & Editing. **Till Bärnighausen:** Conceptualization, Funding acquisition, Investigation, Methodology, Software, Supervision, Writing – Review & Editing. **Ann Blandford:** Conceptualization, Investigation, Methodology, Software, Writing – Review & Editing. **Adrian Dobra:** Conceptualization, Formal analysis, Methodology, Writing – Review & Editing. **Dickman Gareta:** Data curation, Software, Writing – Review & Editing. **Maxime Inghels:** Conceptualization, Data curation, Formal analysis, Methodology, Validation, Writing – Review & Editing. **Hae-Young Kim:** Conceptualization, Data curation, Formal analysis, Methodology, Writing – Original Draft Preparation. **Thulile Mathenjwa:** Conceptualization, Investigation, Methodology, Project Administration, Software, Writing – Review & Editing. **Phillippa Matthews:** Conceptualization, Funding acquisition, Methodology, Software, Writing – Review & Editing. **Nuala McGrath:** Conceptualization, Funding acquisition, Methodology, Software, Writing – Review & Editing. **Jane Seeley:** Conceptualization, Funding acquisition, Methodology, Software, Writing – Review & Editing. **Maryam Shahmanesh:** Conceptualization, Funding acquisition, Methodology, Software, Writing – Review & Editing. **Frank Tanser:** Conceptualization, Funding acquisition, Investigation, Methodology, Software, Supervision, Writing – Review & Editing. **Sally Wyke:** Conceptualization, Funding acquisition, Methodology, Software, Writing – Review & Editing. **H Manisha Yapa:** Conceptualization, Methodology, Writing – Review & Editing. **Thembelihle Zuma:** Conceptualization, Methodology, Writing – Review & Editing.

## Abstract

**Introduction:** Linkage to HIV care remains suboptimal among men. We investigated the effectiveness of a male-targeted HIV-specific decision support app, Empowering People through Informed Choices for HIV (EPIC-HIV), on increasing linkage to HIV care among men in rural South Africa.

**Methods:** Home-Based Intervention to Test and Start (HITS) was a multi-component cluster-randomized controlled trial among 45 communities in uMkhanyakude, KwaZulu-Natal. The development of EPIC-HIV was guided by self-determination theory and human-centered intervention design to increase intrinsic motivation to seek HIV testing and care among men. EPIC-HIV was offered in two stages: EPIC-HIV 1 at the time of home-based HIV counseling and testing (HBHCT), and EPIC-HIV 2 at 1 month after positive HIV diagnosis. Sixteen communities were randomly assigned to the arms to receive EPIC-HIV, and 29 communities to the arms without EPIC-HIV. Among all eligible men, we compared linkage to care (initiation or resumption of antiretroviral therapy after >3 months of care interruption) at local clinics within 1 year of a home visit, which was ascertained from individual clinical records. Intention-to-treat analysis was performed using modified Poisson regression with adjustment for receiving another intervention (i.e., financial incentives) and clustering at the community level. We also conducted a satisfaction survey for EPIC-HIV 2.

**Results:** Among all 13,894 eligible men (i.e., ≥15 years and resident in the 45 communities), 20.7% received HBHCT, resulting in 122 HIV-positive tests. Among these, 54 men linked to care within 1 year after HBHCT. Additionally, of the 13,765 eligible participants who did not receive HBHCT or received HIV-negative results, 301 men linked to care within 1 year. Overall, only 13 men received EPIC-HIV 2. The proportion of linkage to care did not differ in the arms assigned to EPIC-HIV compared to those without EPIC-HIV (adjusted risk ratio=1.05; 95% CI:0.86-1.29). All 13 men who used EPIC-HIV 2 reported the app was acceptable, user-friendly, and useful for getting information on HIV testing and treatment.

**Conclusion:** Reach was low although acceptability and usability of the app was very high among those who engaged with it. Enhanced digital support applications could form part of interventions to increase knowledge of HIV treatment for men.

Clinical Trial Number: ClinicalTrials.gov # NCT03757104

## INTRODUCTION

Despite significant progress in HIV prevention and antiretroviral therapy (ART),^1–8^ timely linkage to care remains a challenge, especially among men in sub-Saharan Africa.^9^ In South Africa, of 8 million people living with HIV (PLWH), 94% of PLWH knew their HIV status, but only 74% of them linked to care and received ART in 2021.^10^ Gaps are particularly significant among men, with only 68% successfully linking to care and receiving ART, whereas nearly 80% of women did so.^10^ Home-based HIV counseling and testing (HBHCT) has been demonstrated as a highly acceptable and effective way to increase HIV testing and diagnosis in sub-Saharan Africa (SSA).^11–18^ However, studies have reported that linkage to HIV care at clinics after HBHCT has been suboptimal, ranging from as low as 14% to 75% ^16,19–22^ In particular, men encounter various barriers such as social stigma, gender norms, or perceptions about the treatment, resulting in delays in seeking treatment until they feel severely ill.^21,23^ Also, men often report the need to prioritize going to work over visiting a clinic for financial reasons.^24^

Linking to and remaining in care for life-long treatment require consistent and internal motivation. Motivational interviewing (MI) is widely used as a person-centered counseling intervention to enhance intrinsic motivation for behavior change.^25^ Studies have demonstrated its effectiveness in increasing adherence to ART^26^ or reducing sexual risky behavior among both HIV-positive and HIV-negative individuals.^27^ It is conceptually based on self-determination theory (SDT), which postulates that meeting three essential psychological needs – autonomy (i.e., having a sense of choice and being the initiator of actions), competence (i.e., feeling capable of achieving desired outcomes), and relatedness (i.e., need to feel connected to others) – enhances individuals’ motivation for behavioral changes and facilitate achieving desired health outcomes over time.^28–31^ However, implementing and delivering MI can be resource-intensive. Counselors need to receive comprehensive and specialized training,^32^ and MI sessions typically lasts about 30-40 minutes per session and multiple sessions might be required to achieve effective behavior change.^33^

Digital technology interventions are increasingly utilized as one of the approaches to address this challenge and support engagement in the HIV care continuum.^34^ While these interventions require infrastructure such as access to phones and data connectivity, they have the potential to be more cost-effective, less labor-intensive, and more equitable to overcome geographical barriers. Several studies have examined the effect of digital technologies to provide counseling and support on improving linkage to care in SSA. In rural Uganda and Kenya, when individuals who tested positive via community-wide HIV testing were offered facilitated linkage to care such as provision of a telephone hotline for enquiries, up to 73% linked to care within 1 year.^35^ On the other hand, less than one-third of individuals newly diagnosed via HBHCT linked to care within 6 months in rural South Africa, despite SMS reminders and nurse-led telephone support for linkage to care.^9^ To our knowledge, no study has explicitly developed a tablet-based male-targeted HIV decision support app to provide tailored counseling based on SDT and examined its impact on linkage to HIV care in SSA.

Home-Based Intervention to Test and Start (HITS) was a multi-component cluster-randomized controlled trial among 45 communities in uMkhanyakude district of KwaZulu-Natal. We previously reported that the probability of uptake of HIV testing increased by more than 50% in the financial incentive arms but did not differ in the arms which received a male-targeted HIV-specific decision support app, Empowering People through Informed Choices for HIV (EPIC-HIV).^36^ In this study, we investigated the impact of EPIC-HIV on increasing HIV-positive diagnosis and linkage to HIV care within 1 year among men in rural South Africa.

## METHODS

### Setting

The trial was nested within AHRI’s Health and Demographic Surveillance System (HDSS) and facilitated linkage to care in the uMkhanyakude district of northern KwaZulu Natal.^37^ Adult (age 15+ years) HIV prevalence in the study area was estimated as 19% in men and 40% in women in 2018.^1^ Since 2003, AHRI has conducted ongoing population-based HIV surveillance within AHRI’s HDSS. The HIV surveillance is annually conducted among all residents a^1^ged ≥15 years to collect data on sexual behavior and general health as well as dried blood spots samples for anonymized HIV testing after obtaining informed consent.^38,39^

### Trial Design

The trial was registered at the National Institute of Health (ClinicalTrials.gov # NCT03757104), and the full trial protocol was published elsewhere.^40^ In brief, between February and December 2018, we implemented a 2x2 factorial cluster-randomized trial among a population of 37,028 residents aged ≥15 years across 45 clusters within AHRI’s ongoing population-based annual HIV testing platform (Figure 1).^40–42^ Two interventions were conditional financial incentives and a male-targeted app-based system for informed decision making, called EPIC-HIV. Over the entire study duration, 8 communities received only financial incentives, 8 communities received only EPIC-HIV, 8 communities received both intervention, and 21 communities received standard of care. Both males and females were eligible to receive financial incentives but only males were eligible to receive EPIC-HIV. Implementation and acceptance of the HITS intervention were evaluated using a process evaluation through post-intervention satisfaction surveys^43,44^ as well as focus group discussion and in-depth interviews among study participants, fieldworkers, and health professionals. The results of the process evaluation are published elsewhere.^45^ Randomization was conducted to ensure balance across the arms using stratified sampling at the community-level based on the HIV incidence among young females aged 15-30 years.

**Figure 1.**
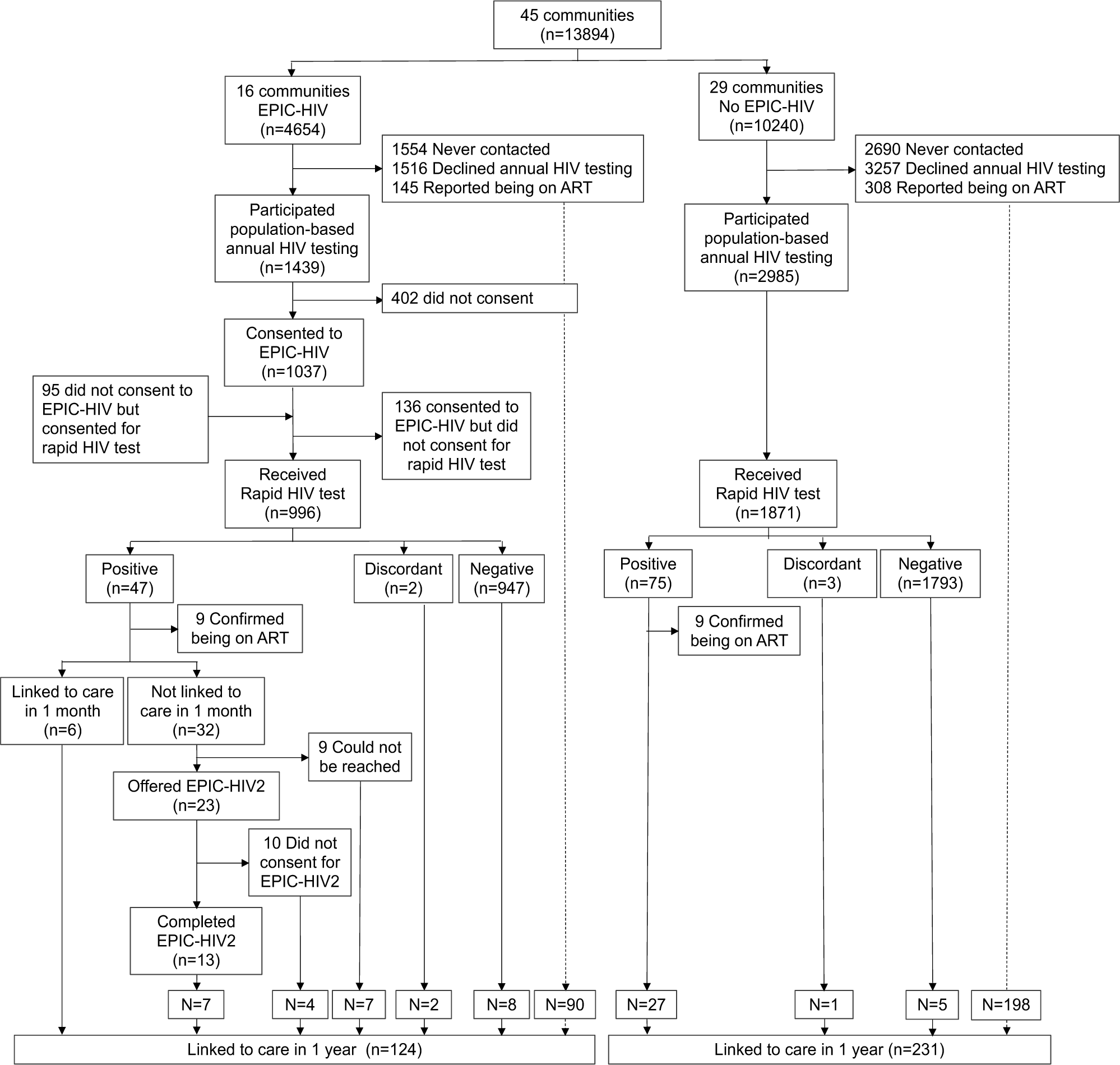
Flow diagram for the HITS cluster-randomized controlled trial and linkage to care within 1 year of a home visit among men. Flow diagram shows individual flow through each stage of the HITS trial in the arms with and without EPIC-HIV. The dashed line indicates linkage to care within 1 year among those who were never contacted, declined annual HIV testing, or did not consent to EPIC-HIV.

**Figure 2.**
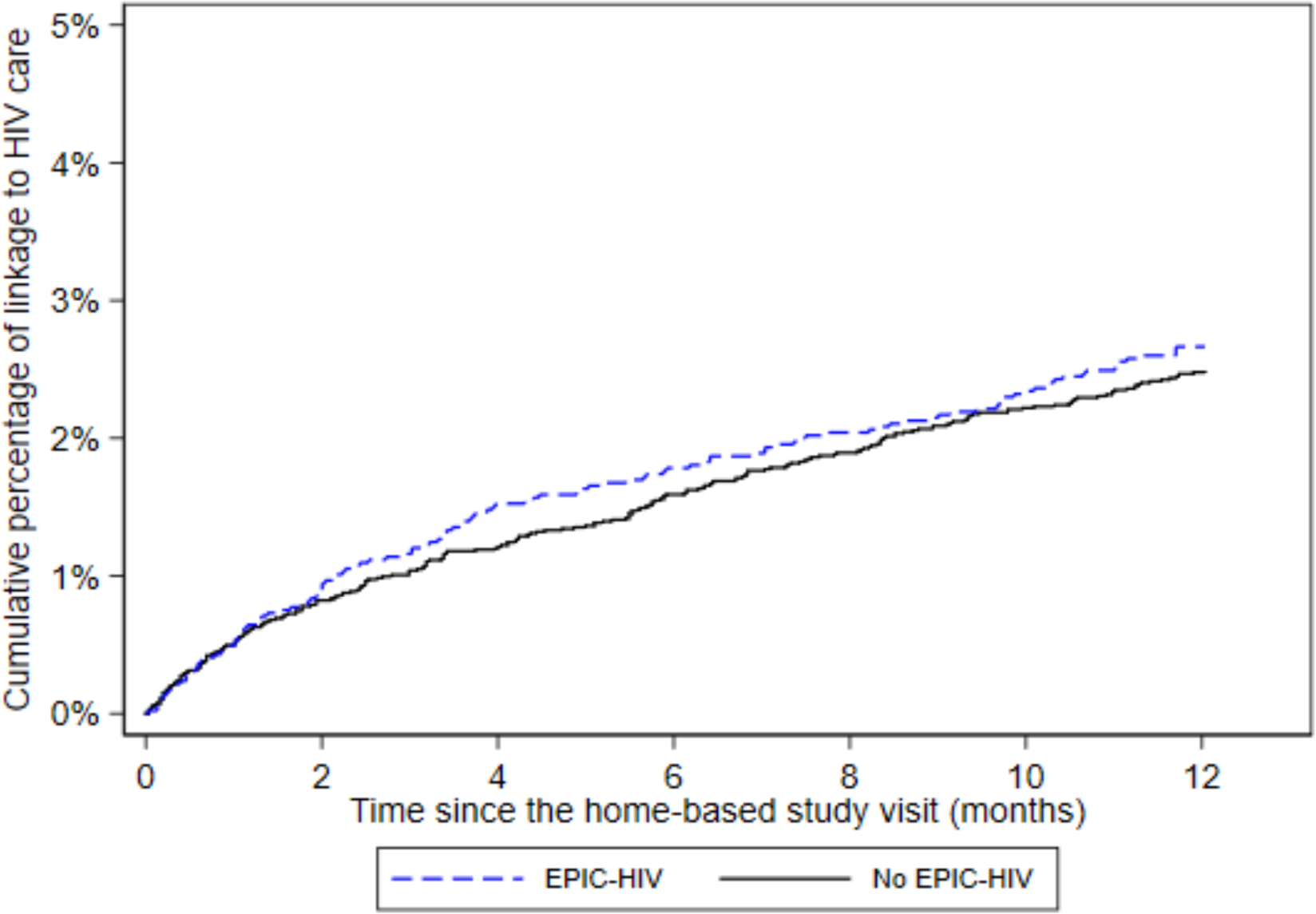
Kaplan Meir curves for linkage to HIV care at clinics for one year after the home-based study visit among men. The solid black lines represent the arms without EPIC-HIV, and the dashed blue lines the arms with EPIC-HIV.

### Standard of care for HBHCT and linkage to care

Since 2017, a rapid point-of-care HIV test with immediate result has been offered by field workers as part of the AHRI population-based annual HIV surveillance. All field workers are trained in HIV counselling and testing in accordance with the South African national guidelines. If individuals are not available at the initial attempt of contact, field workers revisit them two more times in the same week during normal working hours. If they are still not available, a tracking team makes a home visit for three more times in the evenings or weekends before considering them as not being able to contact. Participants testing HIV-positive via HBHCT are encouraged to link to care within 7-10 days of the HIV test date, and are offered a referral slip for an appointment to receive HIV care at one of the 10 local clinics. AHRI has set up an electronic data collection system, called ARHILink, which records clinical information and reasons for visits for all consenting individuals presenting at these clinics including ART initiation and ART follow-up visits. Also, the AHRI surveillance database was linked with TIER.Net, a three-tiered electronic patient management system used in public clinics in South Africa for monitoring and evaluating HIV care and treatment for all patients on ART using personal identifiers and algorithms developed at AHRI. Both AHRILink and TIER.Net were used to confirm the date of ART initiation or resumption of ART care after the study visit among all eligible participants in the trial. Individuals who opt in and consent for facilitated linkage and have not linked to care within two weeks of the HIV test date receive a single Short Message Service (SMS) message as a reminder. If these individuals do not link to care within a further two weeks, a trained nurse contacts them by telephone to discuss any concerns and encourages them to link to care.^9^

### Intervention

The male-targeted HIV-specific decision support application (EPIC-HIV) was developed in two versions (EPIC-HIV 1 and EPIC-HIV 2) and implemented and offered via a tablet. The application development is described in detail elsewhere.^40,44^ In brief, the development was guided by a combination of self-determination theory and human-centered intervention design using a mixture of audio, text, video, photos, and graphics. The conceptual design was first developed in three phases: 1) literature review on barriers and facilitators for HIV care among men; 2) prototype development and iteration drawing on review and self-determination theory, with a specific focus on autonomy and competence, with some aspects of relatedness; and 3) evaluation with target users. Then the prototype of the app was electronically developed and refined for design and usability with potential users. EPIC-HIV 1 has seven characters designed to appeal to different clients. These characters explain how and why they overcame barriers to HIV testing and treatment to provide vicarious experience to clients and to encourage their decision to take a rapid HIV test and link to care if tested HIV-positive. EPIC-HIV 2 has three main modules, which are designed to demonstrate how barriers to seeking HIV treatment can be overcome, focusing on the effectiveness of ART to suppress viral load and transmission to sexual partners. All modules were designed to demonstrate that essential features of masculinity would not be challenged by HIV testing and treatment. They were designed in an interactive narrative form, and participants could freely explore different modules using a private earphone. The interventions were delivered in a two-stage scheme for HIV testing and linkage to care. EPIC-HIV 1 was delivered to men prior to the HIV test offer. If participants did not link to HIV care within a month of a positive HIV test, a study tracker re-visited and offered them EPIC-HIV 2.

In the arms receiving financial incentives, participants were offered a R50 (US ∼$3) food voucher for a local supermarket conditional on their participation in rapid HIV testing. Participants who tested HIV-positive were offered another R50 food voucher if they visited any of the 10 primary health clinics in the AHRI HIV surveillance area to seek HIV treatment within 6 weeks of the positive HIV test date.

### Outcomes

In this study, we report the probability of linkage to care at local clinics within 1 year after home-based visit in the arms which received EPIC-HIV and those which did not receive EPIC-HIV. Linkage to care was defined as ART initiation or resumption and captured using two sources of data collection, TIER.Net and AHRILink. ART initiation was defined as being newly initiated on ART without any prior record of ART initiation. ART resumption was defined as re-initiating ART after >90 days of care interruption as ascertained through AHRILink and/or TIER.Net. We also assessed the acceptability of and satisfaction with EPIC-HIV 2 among males who completed EPIC-HIV 2 using the adapted Client Satisfaction Questionnaire (CSQ-8) with eight questions.^46^ In addition, two questions were added to assess the relevance of the app in the setting and user friendliness. All ten items were asked using a 4-point Likert scale of 1 to 4 per item, where the higher points were associated with agreement or satisfaction (e.g., 3 for mostly satisfied and 4 for very satisfied) and the lower points with disagreement or dissatisfaction (e.g., 1 for mostly dissatisfied and 2 for quite dissatisfied). More details of the satisfaction survey completed for EPIC-HIV 1^47^ and EPIC-HIV 2^44^ have been reported elsewhere.

### Statistical methods

The primary analysis was conducted using the intent-to-treat (ITT) analysis for all men randomized at the community level by the arms with and without EPIC-HIV. We examined the outcome as a binary variable using modified Poisson regression with adjustment for financial incentives and clustering of standard errors at the community level. The time to linkage to care by intervention arms by 1 year was estimated using Kaplan-Meir survival curves. The log rank test was used to compare the difference in the linkage to care. For the satisfaction survey items, the proportions who reported satisfaction or positive agreement (i.e., point 3 or 4 in the 4-point Likert item) were calculated. Analyses were conducted in STATA 15.1 (StataCorp) and R 4.0.3.

### Ethics statement

The study protocols for the AHRI’s population-based HIV testing platform and HITS intervention were approved by the Biomedical Research Ethics Committee of the University of KwaZulu-Natal (BE290/16 and BFC398/16).^40^ Permission for the trial was obtained from the KwaZulu-Natal Department of Health, South Africa. Participation in the HIV surveillance and HITS trial is completely voluntary. Individuals may choose not to answer and/or participate in any component of the HIV surveillance and to withdraw at any time. Written informed consent was sought from individuals aged ≥ 18 years, and parental or guardian consent with child assent for individuals of 15-17 years old were obtained.

## RESULTS

### Participants and recruitment flow

All 15,485 men living in the 45 clusters in the study area were initially considered eligible for the trial. Study participants were enrolled from February 2018 to December 2018 and followed up over 1 year. Of these, 1,591 died or out-migrated. Of the 13,894 remaining eligible men, 4,244 (30.6%) were never contacted (mainly due to absence at the time of HIV testing despite several attempts to follow up), 4,773 (34.4%) chose not to participate in the annual population-based HIV testing in 2018, and 453 (3.3%) reported being on ART, thus resulting in 4,424 (31.8% of the resident population) who participated in the population-based HIV testing (Figure 1). Randomization successfully achieved balance regarding HIV prevalence and sociodemographic variables in EPIC-HIV vs. no EPIC-HIV arms except for the area of residency (Table 1).

**Table 1.** Baseline characteristics of communities and individuals among men in the EPIC-HIV vs. no EPIC-HIV arms.

| Characteristic | EPIC-HIV | No EPIC-HIV |
| --- | --- | --- |
| ***Community-level factor*** | **n=16** | **n=29** |
| HIV prevalence in 2018, % (95% CI)* | 18.5 (15.9-21.3) | 15.0 (12.4-17.9) |
| ***Individual-level factors*** | **n=4654** | **n=9240** |
| Time since last HIV test in the surveillance (years), median (IQR)** | 2.5 (0.6, 6.0) | 2.3 (1.0, 5.3) |
| **Age (years), n (%)** |  |  |
| 15-25 | 1703 (36.6) | 3406 (36.9) |
| 25-35 | 1058 (22.8) | 2145 (23.3) |
| 35-45 | 694 (14.9) | 1474 (16.0) |
| 45-55 | 471 (10.1) | 899 (9.7) |
| ≥55 | 721 (15.5) | 1298 (14.1) |
| **Marital Status, n (%)** |  |  |
| Never married | 1209 (26.0) | 2463 (26.7) |
| Married | 706 (15.2) | 1314 (14.2) |
| Informal Union | 1774 (38.1) | 3610 (39.1) |
| Separated/Divorced/Widowed | 93 (2.0) | 159 (1.7) |
| Don't know/Missing | 872 (18.7) | 1694 (18.3) |
| **Education, n (%)** |  |  |
| No formal education | 1046 (22.5) | 1977 (21.4) |
| Primary (grade 1-7) | 254 (5.5) | 499 (5.4) |
| Secondary+ (≥ grade 8) | 2700 (58) | 5401 (58.5) |
| Don't know/Missing | 654 (14.1) | 1363 (14.8) |
| **Area of residency, n (%)** |  |  |
| Rural | 2644 (56.8) | 5397 (58.4) |
| Peri-urban | 1677 (36.0) | 3129 (33.9) |
| Urban | 333 (7.2) | 714 (7.7) |

\*95% confidence intervals were calculated using the cumulative probabilities of the binomial distribution.
\*\*Data indicates the time since last HIV test among those who ever participated in the annual HIV surveillance, including 2,051 (44.4%) in the EPIC-HIV arms and 4,052 (43.7%) in the arms without EPIC-HIV among men. Abbreviation: IQR, Interquartile Range

### HIV positive diagnosis by intervention arms among men

Among all 13,894 men ≥15 years resident in the 45 communities in 2018, the overall uptake of home-based HIV testing was 21.4% (996/4654) in the EPIC-HIV arms, and 18.3% (1871/10240) in the arms without EPIC-HIV. Of 20.6% men (n=2,867) who received HBHCT, 122 were tested HIV-positive. In ITT analysis, 1.0% (47/4654) in the EPIC-HIV arms and 0.8% (75/9240) in the arms without EPIC-HIV were tested HIV-positive using the rapid HIV tests. The probability of positive HIV diagnosis via HBHCT did not differ in the EPIC-HIV arms, compared to the arms without EPIC-HIV, after adjusting for financial incentives and clustering at community-level (adjusted Risk Ratio [aRR]=1.07, 95% CI: 0.69-1.68, p=0.76).

### Linkage to care at 1 year by intervention arms among men

From individual clinical records ascertained from 17 clinics within the surveillance area, we confirmed that a total of 355 men in the trial initiated ART or resumed care within 1 year of the home visit. First, of a total of 129 who tested HIV-positive (n=122) or discordant or invalid (n=7) via HBHCT, 18 were confirmed to be on ART, 58 had interrupted ART care, and 53 never initiated on ART at the time of HBHCT. Among the 111 men who had interrupted ART care or not on ART, 54 (48.6%) linked to care by 1 year (26/40 [65.0%] in the EPIC-HIV arms and 28/71 [39.4%] in the arms without EPIC-HIV). Second, of 13,765 who did not receive HBHCT or received HIV-negative results via HBHCT, 1625 were confirmed to be on ART, 484 had interrupted care at the time of study visit, 11,656 never initiated on ART at the time of HBHCT. By 1 year after the study visit, 10 people newly initiated ART and 291 resumed care (98/4037 [2.4%] in the EPIC-HIV arms and 203/8103 [2.5%] in the arms without EPIC-HIV). Overall, in the ITT analysis, there was no increase in linkage to care within 1 year among men in the EPIC-HIV arms (2.7%, 124/4654 compared to the arms without EPIC-HIV (2.5%, 231/9240), after adjusting for financial incentives and clustering at community-level (aRR=1.05, 95% CI: 0.86-1.29).

### EPIC-HIV 2 and linkage to care

Among the 47 men tested positive in the EPIC-HIV arms, 32 had not been linked to care by 1 month after home-based study visit, 9 men could not be reached, and 23 were offered EPIC-HIV 2. Of these, 13 men consented and completed EPIC-HIV 2. Of the 10 men who did not consent for EPIC-HIV 2, the main reasons were being busy or at work (40%, n=4), understanding the purpose of the app without the need to engage with the app (30%, n=3), and feeling fine (20%, n=2), or other (10%, n=1). Of those who completed EPIC-HIV 2, everyone (n=13) reported that they were mostly or very satisfied with the app as well as the amount of HIV management information they received through the app, and felt motivated to seek HIV care. Also, everyone assessed the user-friendliness of the app as excellent.

Of those who completed EPIC-HIV 2, 8 men (61.5%) successfully linked to care by 1 year, while 4 men (40%) who did not consent for EPIC-HIV 2 did so.

## DISCUSSION

In this study, we found that the provision of a digital counseling app based on self-determination theory and human-centered intervention design did not significantly improve linkage to care by 1 year in men, despite the fact that the participants found that both EPIC-HIV1 (offered at the time of HIV testing)^47^ and EPIC-HIV 2 (offered at 1 month follow-up after positive HIV diagnosis) were highly acceptable, user-friendly, and useful for getting information on HIV testing and treatment. Also, nearly half of them who completed EPIC-HIV2 did not link to care by 1 year after home-based testing and positive HIV diagnosis.

Several factors may have contributed to the null effect of the intervention. First, the implementation and delivery of the EPIC-HIV app proved to be complex and encountered various challenges. EPIC-HIV was specifically designed to accommodate different learning styles and offer tailored information based on SDT principles, allowing end-users to independently navigate the app without requiring direct support or interaction with a counselor.^44^ However, some participants did not use the earphones or needed assistance from the study staff to navigate the app.^45^ Additionally, due to time constraints for both study staff and participants during household visits, some reported that they did not have sufficient time to explore the app or rushed through navigating the app.^45^ Lastly, the number of new HIV diagnoses, and consequently those who were successfully reached and provided with EPIC-HIV 2 was low in this single round of intervention, which would have reduced the power to detect an intervention effect. Building upon the high acceptability of the app, strategies to increase the reach and dose, such as offering the app through mobile phones or targeting a larger audience, could be considered in future.

Men tend to face higher barriers to accessing care at clinics, including perceptions about the treatment, social stigma or gender norms, as well as other structural barriers.^21,23^ A study in KwaZulu-Natal also reported insufficient time or the need to prioritize going to work as barriers for linking to care among men.^24^ In this study, of those who were not linked to care in 1 month, >60% could not be reached or did not consent for EPIC-HIV 2. The primary reasons included being busy at work or feeling fine, which were similar to the barriers to linkage to care found in other studies.^21,23,24^ We also found that small once-off financial incentives increased the uptake of HIV testing but did not improve linkage to care.^36^ Collectively, these suggest that the once-off counselling app or financial incentives might not be sufficient to overcome these socioeconomic and structural barriers for linkage to care. Future studies could investigate the effect of providing additional support to overcome such structural barriers. For example, in Zimbabwe, when patients diagnosed with HIV at home were escorted to visit a clinic and link to care, more than 85% successfully linked to care within 30 days of HIV diagnosis.^48^ In another study in Uganda, when patients were followed up and offered counseling at home 1 and 2 months after HBHCT, it helped to overcome barriers to HIV care such as stigma and fear of treatment side effects, resulting in successful link to care.^49^ Such interventions could be resource-intensive and may need to be adapted to a specific local setting and context.

Despite the study being conducted as a cluster-randomized clinical trial over 13,000 male participants within the ongoing population-based HIV surveillance, the overall number of individuals who were successfully reached and received EPIC-HIV 1 and EPIC-HIV 2 was low. It is also possible that individuals may have linked to care at other clinics within or outside the subdistrict, or linked to care after our pre-specified time frame. We defined linkage to care as ART initiation or resumption, but it is possible that study participants linked to care without initiating ART and were missed in our analysis.

## CONCLUSION

In the multi-component cluster-randomized clinical trial, we found that the provision of a digital counseling app based on self-determination theory and human-centered intervention design did not significantly increase linkage to care by 1 year among men. The high levels of engagement with the digital decision-support application suggests that enhanced digital support applications might form part of a suite of interventions to increase knowledge of HIV treatment and prevention options for men.

## Data Availability

The de-identified datasets are available upon reasonable request through the Africa Health Research Institute (AHRI) data repository at https://data.ahri.org/index.php/home

## Acknowledgement

We thank community members for their continued support and participation in the Africa Health Research Institute’s Health and Demographic Surveillance System (HDSS), and the AHRI population, clinical, laboratory and data management teams.

